# SARS-CoV-2 wastewater surveillance in Germany: long-term PCR monitoring, suitability of primer/probe combinations and biomarker stability

**DOI:** 10.1101/2021.09.16.21263575

**Authors:** Johannes Ho, Claudia Stange, Rabea Suhrborg, Christian Wurzbacher, Jörg E. Drewes, Andreas Tiehm

## Abstract

In recent months, wastewater-based epidemiology (WBE) has been shown to be an important tool for early detection of SARS-CoV-2 circulation in the population. In this study, a detection methodology for SARS-CoV-2 RNA (wild-type and variants of concern) in wastewater was developed based on the detection of different target genes (E and ORF1ab) by PEG precipitation and digital droplet PCR. This methodology was used to determine the SARS-CoV-2 concentration and the proportion of N501Y mutation in raw sewage of the wastewater treatment plant of the city of Karlsruhe in southwestern Germany over a period of 1 year (June 2020 to July 2021). Comparison of SARS-CoV-2 concentrations with reported COVID-19 cases in the catchment area showed a significant correlation. Viral RNA titre trends appeared more than 12 days earlier than clinical data, demonstrating the potential of wastewater-based epidemiology as an early warning system. Parallel PCR analysis using seven primer and probe systems revealed similar gene copy numbers with E, ORF, RdRP2 and NSP9 assays. RdPP1 and NSP3 generally resulted in lower copy numbers, and in particular for N1 there was low correlation with the other assays due to outliers. The occurrence of the N501Y mutation in the wastewater of Karlsruhe was consistent with the occurrence of the alpha-variant (B.1.1.7) in the corresponding individual clinical tests. In batch experiments SARS-CoV-2 RNA was stable for several days under anaerobic conditions, but the copy numbers decreased rapidly in the presence of dissolved oxygen. Overall, this study shows that wastewater-based epidemiology is a sensitive and robust approach to detect trends in the spread of SARS-CoV-2 at an early stage, contributing to successful pandemic management.

## 1. Introduction

In the current pandemic situation, wastewater-based epidemiology (WBE) is considered an important tool to estimate SARS-CoV-2 prevalence, genetic diversity and geographical distribution (European Commission, 2021; Kitajima et al., 2020). Sewage systems provide a feasible approach to survey faecal viruses across an entire region (Hart and Halden, 2020; Hill et al., 2021), even if asymptomatic courses of infection occur (Qi et al., 2018) and at low frequency of clinical diagnostic testing (Lodder and de Roda Husman, 2020). Therefore, wastewater surveillance can provide an alternative method to detect the spread of infections in different areas -especially for regions with limited diagnostic capacity and without a functioning reporting system, such as developing countries (Hart and Halden, 2020; Kitajima et al., 2020). In addition, wastewater surveillance can help detect variations in circulating strains, allowing comparisons among regions and an assessment of the evolution of the viral genome over time (Bisseux et al., 2018). Moreover, wastewater-based surveillance can serve as an early warning system (Chavarria-Miró et al., 2021; Hill et al., 2021; Kitajima et al., 2020). In light of the current pandemic, this approach could determine whether new SARS-CoV-2 infections have occurred in a community or whether the number of infected people decreases after measures in the affected population have been taken (e.g., lockdown or social distancing). Recent studies have shown the overall benefit of WBE for SARS-CoV-2 monitoring (Agrawal et al., 2021a; Medema et al., 2020; Rossmann et al., 2021b; Rossmann et al., 2021a; Westhaus et al., 2021).

For WBE purposes, SARS-CoV-2 is detected in wastewater using PCR-based methods. Multiple sets of PCR primers and probes have been published, targeting different locations of the genome such as the S, N and E gene (Cervantes-Avilés et al., 2021; Kitajima et al., 2020). However, there is a lack of data with respect to the reliability of primers/probes and genes at different positions of the genome, thus hampering the quantitative comparison of results.

Recently, predominantly RT-qPCR methods have been published. However, as an alternative to quantitative real-time PCR, digital droplet PCR (ddPCR) can be used for wastewater monitoring (Cervantes-Avilés et al., 2021), which is a quantitative PCR method allowing an absolute quantification of DNA or RNA. By using ddPCR, a higher sensitivity can be achieved compared to qPCR since PCR inhibitors and the competition of background DNA and target molecules play a negligible role (Rački et al., 2014).

The genome of the SARS-CoV-2 virus is subject to a high mutation rate. Mutations are distributed across the genome and either have no effect on the phenotype (silent mutation) or can lead to altered infectivity and pathogenicity (Mercatelli and Giorgi, 2020). At the end of 2020 and the beginning of 2021, variants (B.1.1.7 [alpha], B.1.351 [beta] and P.1 [gamma]) with increased transmission and clinical importance appeared (Sandoval Torrientes et al., 2021). These SARS-CoV-2 variants were classified as variants of concern (VoC). All three VoCs have the mutation A23063T, also named N501Y, in common, which is involved in the receptor-binding mechanism and may have clinical impacts (Makowski et al., 2021). For the single base mutation, a primer/probe set that allows the specific detection of the mutation has been described (Heijnen et al., 2021; Korukluoglu et al., 2021). The detection of mutations of SARS-CoV-2 in wastewater additionally reveals a more detailed view of the infection process and the local spread of virus strains.

The SARS-CoV-2 RNA is excreted with the faeces of infected patients (Bogler et al., 2020). This viral RNA reaches the wastewater treatment plant (WWTP) via the sewer system, where composite samples for wastewater monitoring are usually taken from the influent. However, relatively little is known about the stability of SARS-CoV-2 RNA in sewer systems (Hart and Halden, 2020). In general, stability is strongly influenced by various environmental factors such as temperature, pH, biological activity or solid content (Foladori et al., 2020). In laboratory experiments, significantly higher reductions were determined at higher temperatures (Ahmed et al., 2020; Gundy et al., 2009). Coronaviruses are enveloped viruses, and an intact envelope is important for virus pathogenicity (Mandala et al., 2020). Overall, there are indications that enveloped viruses are less resistant to environmental conditions and inactivation compared to non-enveloped viruses such as adeno-or noroviruses (Gundy et al., 2009; Ye et al., 2016). The virus envelope is highly sensitive to chemical and physical conditions (e.g., pH, lipid solvents, disinfectants) (Mohan et al., 2021; Scheller et al., 2020). In contrast, the capsid protein is probably less subjected to lipid solvents, temperature and pH changes, although so far, little information is available on its stability.

In this context, the aims of this study were (i) to establish a robust wastewater monitoring procedure including the evaluation of different target genes and the use of digital droplet PCR, (ii) to apply the WBE approach in southern Germany, (iii) to determine the distribution of SARS-CoV-2 variants and (iv) to investigate the stability of its genome in wastewater samples.

## 2. Material and Methods

### 2.1 Study sites, sample collection and storage

Long-term monitoring was performed between June 2020 and July 2021 at the WWTP of the city of Karlsruhe. Karlsruhe is the second-largest city of the German federal state Baden-Württemberg and located in southwestern Germany. The WWTP of Karlsruhe has a capacity of 850,000 population equivalents and treats the sewage of about 370,000 people.

We collected 24-h-composite samples (from 0 to 12 pm) twice a week using a flow-proportional autosampler. The collected samples were homogenised using the dispersing and homogenizing tool MiniBatch D-9 (MICCRA GmbH) and transported to the laboratory at 4°C. Upon arrival, the samples were concentrated and analysed according to the method described below. In total, 89 samples were taken in the study area as duplicates. Wastewater samples from other sites in southern Germany (Leonberg, Baden-Wuerttemberg and Berchtesgadener Land, Bavaria) were also used for the evaluation of different target genes (802 samples in total).

### 2.2 Sample processing

#### 2.2.1 Concentration

For sample concentration, a volume of 45 mL of wastewater sample was centrifuged (30 min; 5,000 g) to settle larger particles. The supernatant was than concentrated using PEG/NaCl precipitation as described by Wu et al. (2020), with slight modifications. Briefly, 10% (w/v) PEG 8000 (Carl Roth) and 2.25% NaCl (w/v) (Carl Roth) were added to the sample and mixed. The mixture was agitated on a shaking incubator at 120 rpm for 2 h on ice, and subsequently, samples were centrifuged at 20,000 g for 2 h at 4°C. The supernatant was discarded, and the pellet was resuspended in 600 μL PCR-grade water (ThermoFisher Scientific). For all samples two independent replicates were analysed.

#### 2.2.2 Nucleic acid extraction

Nucleic acids were extracted directly after concentration using the innuPrep Virus DNA/RNA Kit – IPC16 (Analytik Jena GmbH) as described by the manufacturer (protocol for isolation from 600 µL cell-free body fluid). Extracted nucleic acids were eluted in a final volume of 100 µL and analysed immediately or stored at -80°C.

#### 2.2.3 PCR detection

The SARS-CoV-2-specific sequences were quantified using one-step RT-ddPCR. For the PCR analyses, seven primers/probe sets were compared (Table 1). All wastewater samples were analysed using two duplex PCR assays with the primers and probe sets 1–4. Additional investigations of selected samples for the evaluation of alternative primers and probes (set 5-7) and the detection of the N501Y mutation and wildtype (sets 8 and 9) were carried out. All primers and probes were obtained from ThermoFisher Scientific.

**Table 1:**
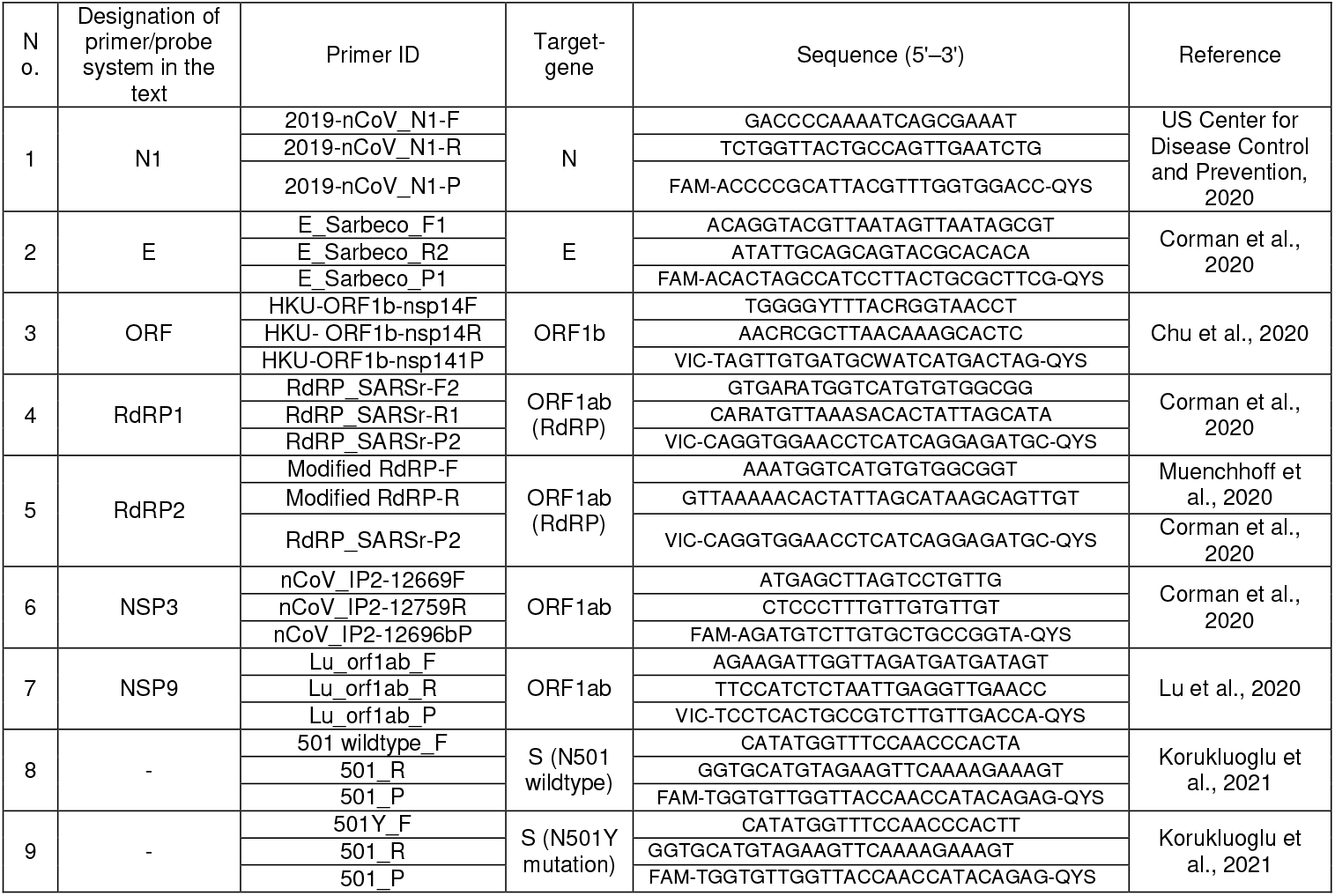
Primers and probes used for ddPCR analysis

The ddPCR was performed on the QX200 Droplet Digital PCR System (BioRad) using the One-step RT-ddPCR Advanced Kit for Probes (BioRad). Reactions were set up in a final volume of 20 µL, following the manufacturer’s instructions, using 3 µL of nucleic acid extract. The reaction mixture consisted of One-step RT-ddPCR Supermix (BioRad), 20 units/µL reverse transcriptase (BioRad), 15 mM DTT (BioRad), 900 nM primer (forward and reverse), 250 nM probe and RNase-free water. The mixture was combined with 70 μL droplet generation oil in the Droplet Generator (BioRad), and the resulting droplets were transferred to a 96-well plate for PCR cycling. Cycling conditions were as follows: 60 min reverse transcription at 42°C (1 cycle), 10 min enzyme activation at 95°C (1 cycle), 30 s denaturation at 95°C/1 min annealing/extension cycle at 55°C (40 cycles), 10 min enzyme deactivation at 98°C (1 cycle) and a hold step at 4°C until reading on the QX100 droplet reader (BioRad). Data analysis was performed with the QuantaSoft software (BioRad). No-template control and a positive control (synthetic RNA, 4BLqSARS-CoV-2 RNA, 4base lab AG) were included in each ddPCR assay. QuantaSoft and QuantaSoft Analysis Pro (BioRad) were used to manually threshold and export the data. Concentrations per reaction were converted to copies per mL of wastewater.

Previous reports showed an actual detection limit of 3 copies per reaction for the ddPCR (Alteri et al., 2020). In this case, the detection limit also represents the limit of quantification. Based on the initial volume of wastewater and the volume of RNA extract used in the PCR reaction, a detection limit of 2.5 gene copies per mL of wastewater was calculated.

### 2.3 COVID-19 cases in the model area

Epidemiological data on COVID-19 cases in the studied area were retrieved from the daily report on COVID-19 infections in the city and districts of Karlsruhe (https://corona.karlsruhe.de/aktuelle-fallzahlen), which is based on official data of the Federal Robert Koch Institute in charge of public health surveillance. In addition, the Association of Accredited Laboratories in Medicine (ALM e.V.) submit the number of variant-specific PCR examinations for Baden-Württemberg on a weekly basis; reports containing this data are published regularly by the State Health Office of Baden-Württemberg.

### 2.4 Statistical analysis

Basic mathematical calculations (mean, median, sliding average) were performed in Microsoft Excel (14.0.7212.5). More complex statistics were calculated in Python (3.7.6) with the SciPy library (1.6.2) according to Virtanen et al. (2020). Bonferroni-correction was applied to all p values by multiplication with the number of samples (n).

### 2.5 Biomarker stability tests

Batch experiments were established in 1-L glass bottles using untreated wastewater from the WWTP of Karlsruhe. The 24-hr composite sample of the previous day (4 L) was transported to the laboratory and directly used for the experiment. Overall, six test bottles containing 600 mL of wastewater were set up. Three bottles with wastewater were purged with ambient air using aquarium pumps, and the other three bottles were purged with nitrogen gas for ten minutes and then sealed immediately. The bottles were incubated at room temperature. Samples were taken to determine dissolved oxygen (DO) and pH using a WTW meter (36020 IDS) with optical sensor (FDO 925) and pH electrode (SenTrix) from Xylem Analytics. After each sampling the wastewater was purged again with ambient air or nitrogen for five minutes. The PCR samples (40 ml) were processed using the Vac-Man Laboratory Vacuum Manifold (Promega) and the Maxwell^®^ RSC Enviro Wastewater TNA Kit (Promega).

## 3. Results and discussion

### 3.1 Suitability of four different primer and probe systems

The ddPCR data for a total of 802 field samples, including results for four primer and probe sets targeting N, E, ORF and RdRP genes (2019-nCoV-N1, E_Sarbeco, HKU-ORF1b-nsp14, RdRP_SARS; hereinafter referred to as N1, E, ORF and RdRP1), were compared (Fig. 1). The standard deviation of independent replicates was 26%. Using N1, 94% of the samples showed higher values than the detection limit. For other primer and probe combinations the proportion of samples showing values higher than the detection limit were considerably lower (62% for E, 54% for ORF and 20% for RdRP1). A similar trend was observed for gene copy numbers, with highest values for N1 and lowest one for RdRP1. Median values were 27.5, 11.8, 11.5 and 8.5 gene copies per mL for N1, E, ORF and RdRP1, respectively.

**Fig. 1.**
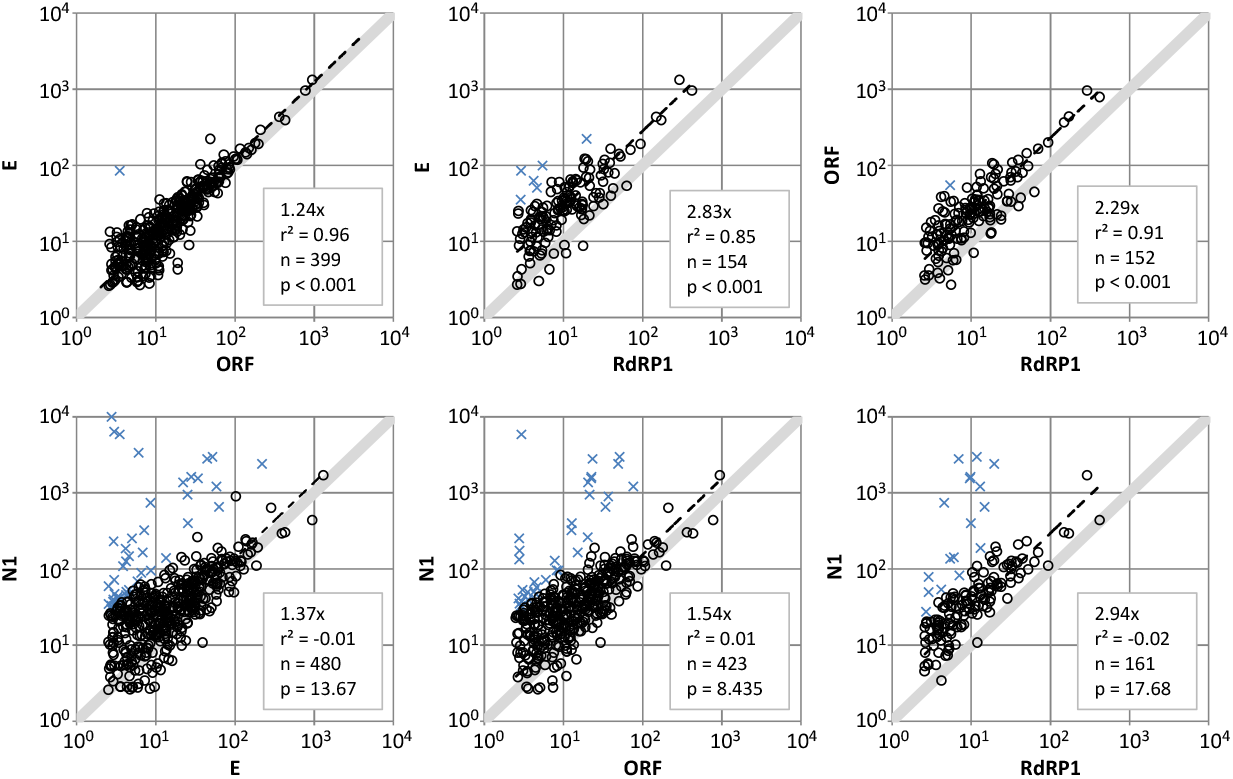
Linear regression of four different primer/probe systems for the analysis of 802 samples. Charts show gene copies per mL wastewater. Values with ≥ tenfold deviation are shown as X. Trend lines were generated using all values.

In particular for N1, we observed several outliers. In some cases, the N1 values were more than 2 log_10_ levels higher than the values obtained by the other assays. Linear regression between the data (Fig. 1) did show low correlation coefficients or no significance (p > 0.05) for all combinations with the N1 primer/probe set. The E and ORF correlated well, with ORF giving slightly lower values as compared to E (slope = 1.24 at R^2^ = 0.96, p < 0.001 and n = 399). Both E and ORF gave higher slope values of 2.83 and 2.29 compared to RdRP1, indicating a noticeable difference. To exclude internal errors for the N1 assay, these extreme values were confirmed by additional replicates.

The copy numbers of the four tested assays differed showing the following order: N1 > E ≈ ORF RdRP1. It was hypothesised that the differences in the gene copy numbers may be due to a non-optimal primer design or the localisation and stability of the target genes on the SARS-CoV-2 genome. The second hypothesis was supported by the preliminary observation that the gene copy numbers decreased towards the 5’ end indicating a preferential degradation of the SARS-CoV-2 genome from the 5’end. This hypothesis was tested with additional primers on the same sample set (see Section 3.3).

For the same primers, differences between gene copy numbers or Ct values have also been reported previously. For example, Vogels et al. (2020) observed similar Ct values for N1, E and ORF but higher Ct values for RdRP1 in clinical samples. Similar findings with N1, E and RdRP1 have been reported by Muenchhoff et al. (2020) for stool samples and for E and RdRP1 in wastewater samples (Bertrand et al., 2021). Higher copy numbers (corresponding to lower Ct values) for the N1 primers in wastewater have been reported by Gerrity et al. (2021) in comparison to N2, E and ORF, by Pérez-Cataluña et al. (2021) in comparison to N2 and E, by Randazzo et al. (2020) for N2 and N3 as well as by Fernandez-Cassi et al. (2021) and Peccia et al. (2020) for N2. Due to low values using RdRP1 and the high outliers using N1, WBE data reporting in our study was performed using only E and ORF. Similar decisions have been made by Gonzalez et al. (2020) and Westhaus et al. (2021); both studies excluded N1 for wastewater monitoring. Considering the observed effects, initial comparison of detection methods and the parallel detection of at least two SARS-CoV-2-specific genes is recommended to assure reliable WBE.

### 3.2 Results of the WBE for the city of Karlsruhe

For the investigated WWTP, 89 samples were taken during the 1-year survey from June 2020 to July 2021; this period included both the second and third waves of COVID-19 infections in Germany. For each sample, two independent replicates were analysed using E and ORF assays. An average SARS-CoV-2 RNA concentration was calculated from the results of the two replicates and the gene copy numbers for E and ORF. Figure 2 displays infection numbers and SARS-CoV-2 RNA concentrations (sliding average of three) over time. Both data curves show a similar but time-delayed trend (Fig. 2, A). By shifting the individual diagnostic testing data set forward, the correlation coefficient increased to a maximum of 0.89 (p < 0.001) at 16 days (Fig. 3 and Fig. S1 and S2 in Supplementary Information). With this time shift, case numbers and ddPCR data matched at increasing and decreasing phases of the 2^nd^ and 3^rd^ waves.

**Fig. 2.**
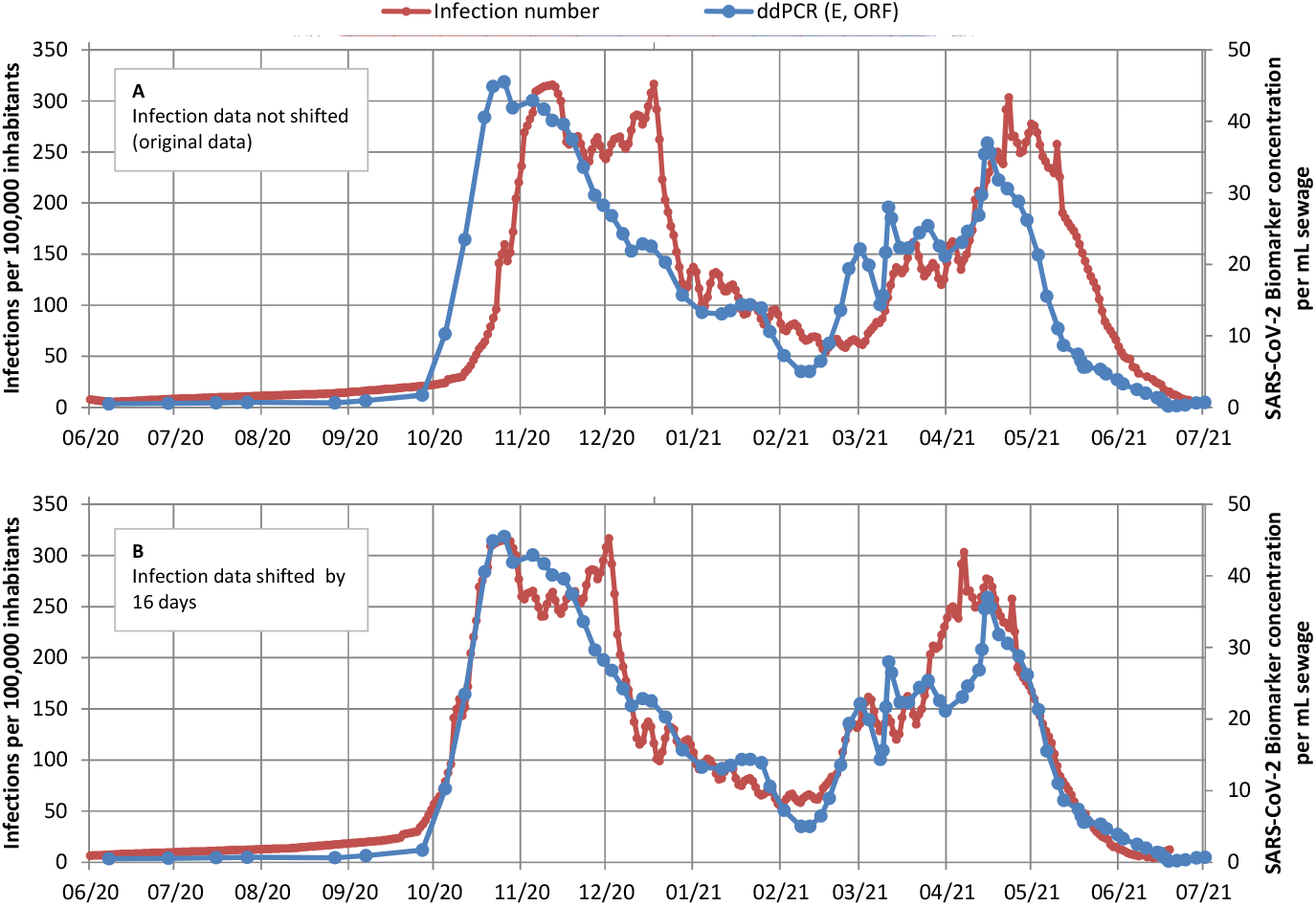
(A) Results of wastewater monitoring and infection numbers and (B) time-shifted infection numbers and biomarker concentrations for the study area.

**Fig. 3.**
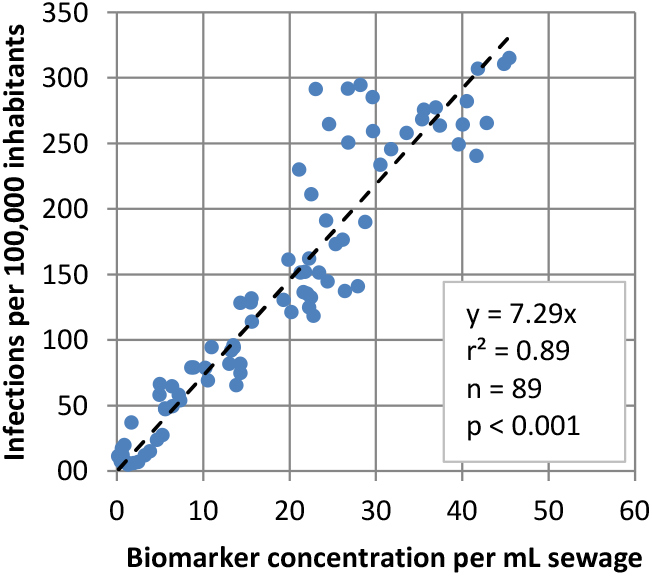
Correlation of infection numbers and ddPCR values with a time shift of 16 days.

The observed time gain of 16 days for WBE in our study is higher than the values reported in previous studies. Time shifts in WBE of 2–4 days (Hillary et al., 2021; Nemudryi et al., 2020), 6–8 days (Peccia et al., 2020), 7 days (Medema et al., 2020), 8 days (Hamouda et al., 2021; Wurtzer et al., 2020), 11 days (Róka et al., 2021) and 10–14 days (Agrawal et al., 2021a) have been reported. In some studies, no correlation could be found due to PCR noise or low case numbers (D’Aoust et al., 2021; Westhaus et al., 2021).

These varying findings can be explained with the high number of variables in diagnostic individual testing: availability of rapid antigen tests, the speed of clinical tests (availability, workload of labs, testing strategy), positive testing rate and the speed of reporting. These factors might change throughout time and differ among SARS-CoV-2 waves (e.g., lockdowns or applied testing strategies). Wastewater monitoring has the advantage that the data for the catchment area can be available with a single measurement within 48 hours. In summary, good correlation of case numbers and SARS-CoV-2 gene copy numbers could be achieved with the detection of the E and ORF genes. The shapes of the E and ORF curves reflect the observed development of COVID-19 cases.

Based on the correlation and a methodological detection limit of 2.5 genomic copies per mL sewage by ddPCR, the theoretical limit of detection for the wastewater monitoring is approx. 20 infections per 100,000 inhabitants. The results of another German study indicated that RT-qPCR can detect 50 acute infected persons per 100,000 inhabitants in dry-weather periods (Westhaus et al., 2021). In our study one genomic copy per mL is equivalent to 7 active cases per 100,000 inhabitants. However, the case/gene copy number factor is hard to compare with literature, as parameters often differ (e.g., cumulative cases (Medema et al., 2020) or gene copies per day (Agrawal et al., 2021a)). Furthermore, these values are highly specific for a sewer system and depend on numerous boundary conditions, such as characteristics of the sewer system. Therefore, SARS-CoV-2 copy number/number of infections factors cannot be transferred directly to other catchment areas.

### 3.3 Extended evaluation of primer and probe systems

The results for the primers/probes N1, E, ORF and RdRP1 exhibited the highest gene copy numbers for N1 and the lowest one for RdRP1. Based on these results, it could be hypothesised that the SARS-CoV-2 genome degradation might preferably start from the 5’ end. In general, viral RNA can be degraded enzymatically via endonucleases, 3’-to-5’ exonucleases and 5’-to-3’ exonucleases (Houseley and Tollervey, 2009). However, viruses have developed specific mechanisms to counteract this degradation. These include various 5’ end modifications, as well as processes of mimicking cellular RNAs via ‘cap snatching’ or even direct blocking of receptor proteins, thus preventing recognition (Dickson and Wilusz, 2011; Markiewicz et al., 2021). The genome of coronaviruses has a 5′-terminal cap structure and a poly(A) sequence at the 3′-end (Kim et al., 2020). To our knowledge, information about the specific enzymatic degradation processes of SARS-CoV-2 RNA in wastewater is not available yet.

To test the hypothesis of the preferential 5’ end degradation, two additional genes located more closely to the 5’ end, namely NSP3 and NSP9, and an alternative primer for the RdRP gene (hereinafter referred to as RdRP2) were investigated with a set of 64 samples (Fig. 4). For this set, samples with high, medium and low copy numbers were selected. High N1 outliers were excluded.

**Fig. 4.**
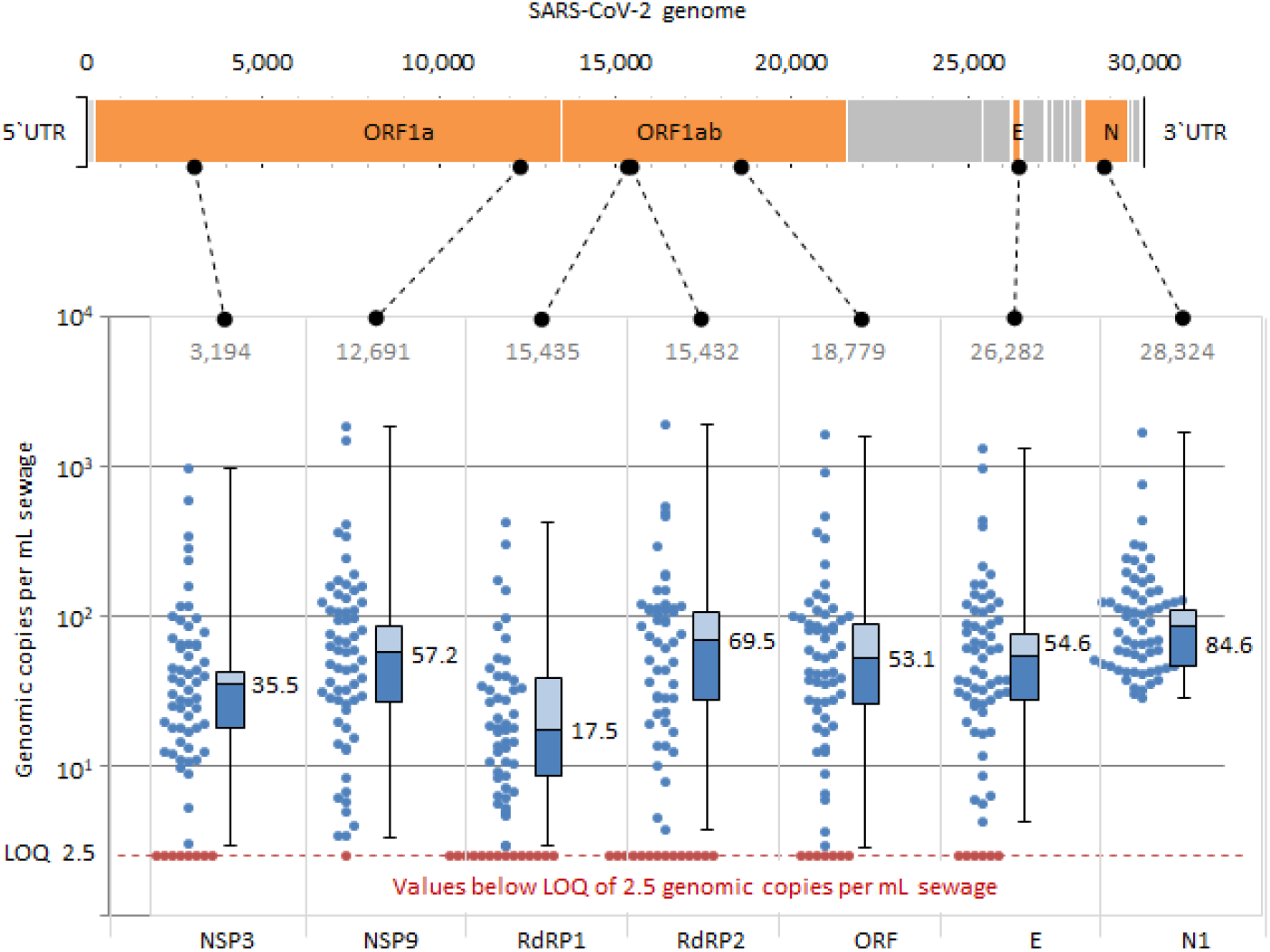
The ddPCR results of 64 samples with seven different primer and probe sets. Dots represent single values. Bars represent 25 and 75%-intervals, numbers represent the medians. SARS-CoV-2 gene and primer positions according to the reference genome NC_045512.2 (Wu et al., 2020).

Generally, the observed median gene copy numbers differed between 17.5 (RdRP1) and 84.6 copies per mL (N1). The medians followed the order N1, RdRP2, NSP9, E, ORF, NSP3, RdRP1. The E, ORF, RdRP1, RdRP2, NSP3 and NSP9 primer/probe sets showed a good correlation with coefficients up to 1.00 (p < 0.05) (Fig. S3 in Supplementary Information). All correlation coefficients with the N1 assay were generally lower and ranged from -0.24 (RdRP2) to 0.74 (ORF). Comparing the slopes of the linear regression, nearly all significant correlations, except those with RdRP1, ranged from 0.5x to 2.0x (n = 46–58, p < 0.05). Generally, RdRP1 provided considerably lower values than the other genes (0.23x-0.46x, n = 50–47, p < 0.05).

Based on this analysis, we can conclude the following: (i) due to the outliers observed with N1 and the low gene copy numbers using RdRP1 and NSP3, these primer/probe sets are not recommended for wastewater surveillance, (ii) E, ORF, RdRP2 and NSP9 worked well for monitoring using ddPCR and provided comparable gene copy numbers (average of deviations: 14%) and (iii) the hypothesis of the 5’-directed degradation of the SARS-CoV-2 genome was rejected.

The effect of deviating between the different PCR-assays might be explained with variations in PCR efficiency and secondary structures of target regions in the viruses. Primers and probes did not show any mismatches in comparison with the corresponding target sequences (e.g., SARS-CoV-2 reference sequence NC_045512.2), except for the RdRP1 reverse primer with a single nucleotide mismatch in the middle. This mismatch could lead to lower copy numbers (Vogels et al., 2020); however, single mismatches not in close 3’proximity are not that critical (Lefever et al., 2013).

Overall, we strongly recommend the application of different primers and probes. Such an approach will assure reliable and robust results and prevent underestimation of results due to newly occurring primer binding site mutations. Additionally, multiple targets will ensure comparability with earlier results in the case a primer has to be exchanged. A good example for this is the N3 forward primer published by the CDC, with a mutated nucleotide position in about 4% of observed sequences (Vogels et al., 2020).

### 3.4 Spread of VoC in the model area

We also quantified VoC of sewage samples in the sewershed of the city of Karlsruhe. The proportion of N501Y to wildtype sequences was compared with publicly available data from the diagnostic tests of infected persons. The proportion of the N501Y mutation in wastewater follows the same course as the trend seen for infected persons (Fig. 5).

**Fig. 5.**
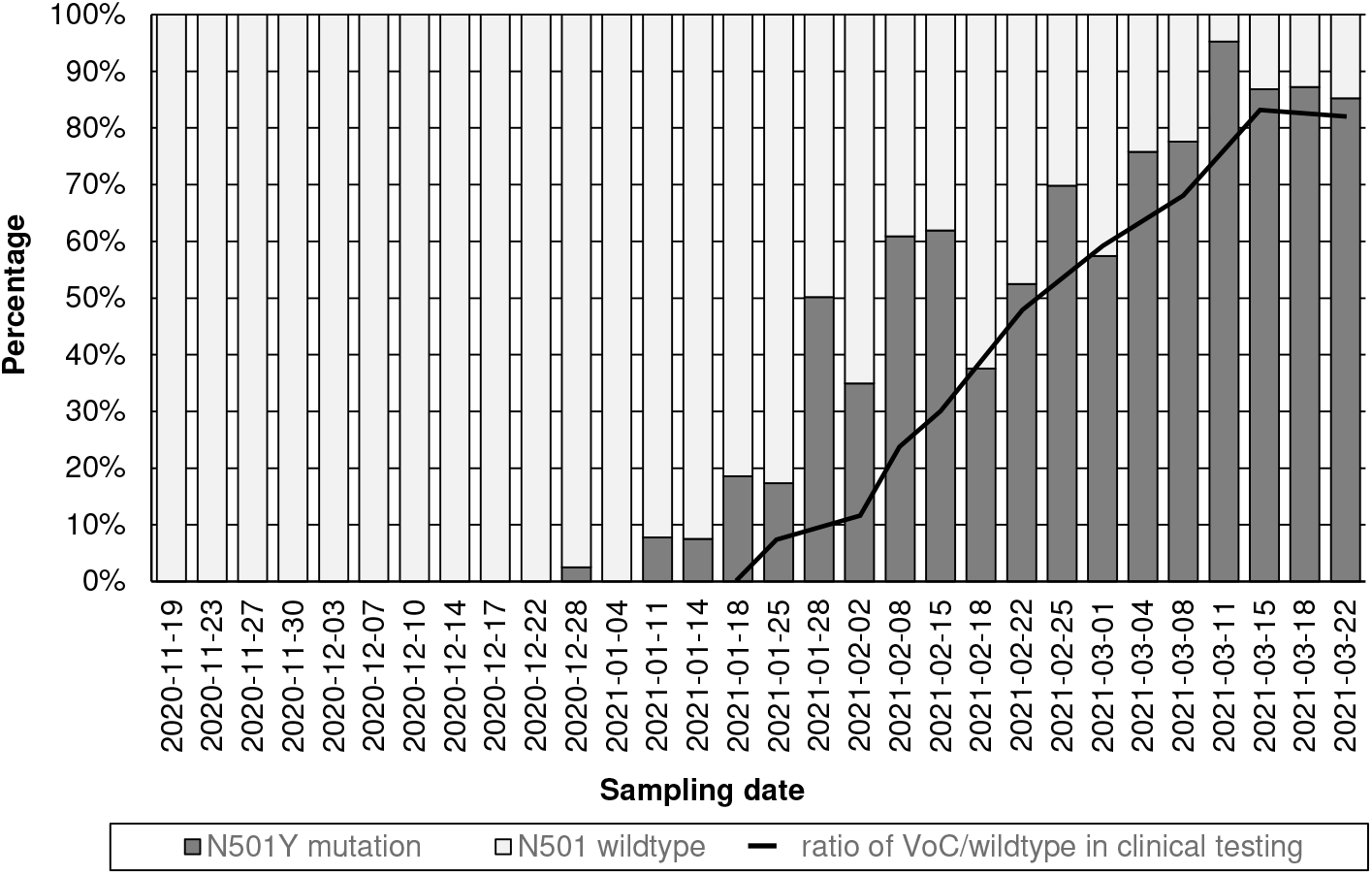
Concentration of N501Y mutation divided by the total gene copy numbers of the S-gene (ratio of wildtype and N501Y mutation) in wastewater, and ratios of variants of concern (VoC) in new COVID-19 patients in Baden-Württemberg by variant-specific PCR according to the Association of Accredited Laboratories in Medicine (ALM e.V).

The earliest detection of N501Y was made in the wastewater sample of December 28, 2020. The proportion of N501Y gradually increased to 97% on March 11, 2021, and then settled at around 86%. In December 2020, the state of Baden-Württemberg reported the detection of the alpha variant (B.1.1.7) for the first time, corresponding to the first detection of N501Y in wastewater from Karlsruhe. After the first detections of variants containing the N501Y mutation, the increase followed a trend that is comparable with the increase of N501Y-containing variants in patients in Baden-Wuerttemberg.

Analogous to a Dutch study (Heijnen et al., 2021), the results of this study show the applicability of RT-ddPCR to detect the spread of relevant mutations of SARS-CoV-2 in the population. Effluent monitoring of mutations by RT-ddPCR is a rapid and efficient method to detect the occurrence of VoC on a community level and could also be used as an early warning system for SARS-CoV-2 variants. In some studies, next-generation sequencing (NGS) techniques from the clinical sector have also been used to detect SARS-CoV-2 mutations (Agrawal et al., 2021b; Crits-Christoph et al., 2021; Izquierdo-Lara et al., 2021). However, these techniques require deep sequencing and extensive knowledge on bioinformatics-supported interpretation of the results to determine the presence of mutations associated with VoC (Heijnen et al., 2021). Furthermore, NGS results are less quantitative than RT-ddPCR results and usually cannot be generated in a timely manner. This temporal aspect is of particular relevance with regard to an early warning function of wastewater monitoring. The advantage of NGS is the generation of comprehensive information on the existing spectrum of mutations in a sample. Overall, RT-ddPCR and NGS are complementary methods that provide important information on the distribution of N501Y and other relevant mutations at the community level through the analysis of RNA extracts from wastewater samples.

### 3.5 Oxygen-dependent decay of SARS-CoV-2 RNA

Depending on flow velocity, temperature, dissolved organic carbon, and other boundary conditions, the DO concentration in the wastewater of the sewer system can vary. For this reason, the decay of SARS-CoV-2 RNA was investigated under aerobic and anaerobic conditions.

At the beginning of the experiment, DO contents of 2.8 mg/L and SARS-CoV-2 RNA concentrations between 95 and 112 gene copies per mL were determined (Fig. 6). In the bottles aerated with atmospheric oxygen, the DO increased up to 8.5 mg/L while the concentration of SARS-CoV-2 RNA decreased by 1 log_10_ level within 24 h. After 4 days the copy numbers were below the detection limit. In the nitrogen treated bottle, the DO concentration decreased below 0.1 mg/L and viral RNA decreased by about 0.3 log_10_ levels (52%) within one day. Subsequently, the concentrations of SARS-CoV-2 biomarkers remained stable. The different target sequences (ORF, E and RdRP2, NSP3) provided comparable gene copy numbers (maximum standard deviation: 26.9 gene copies per mL; Fig. S4). Overall, the results indicate that the DO concentration can affect the fate of SARS-CoV-2 RNA in wastewater. In addition, the batch experiments demonstrated similar degradation rates of four genes localised at different positions throughout the SARS-CoV-2 genome, and thus also did not indicate any directional degradation of the genome (Section 3.3).

**Fig. 6.**
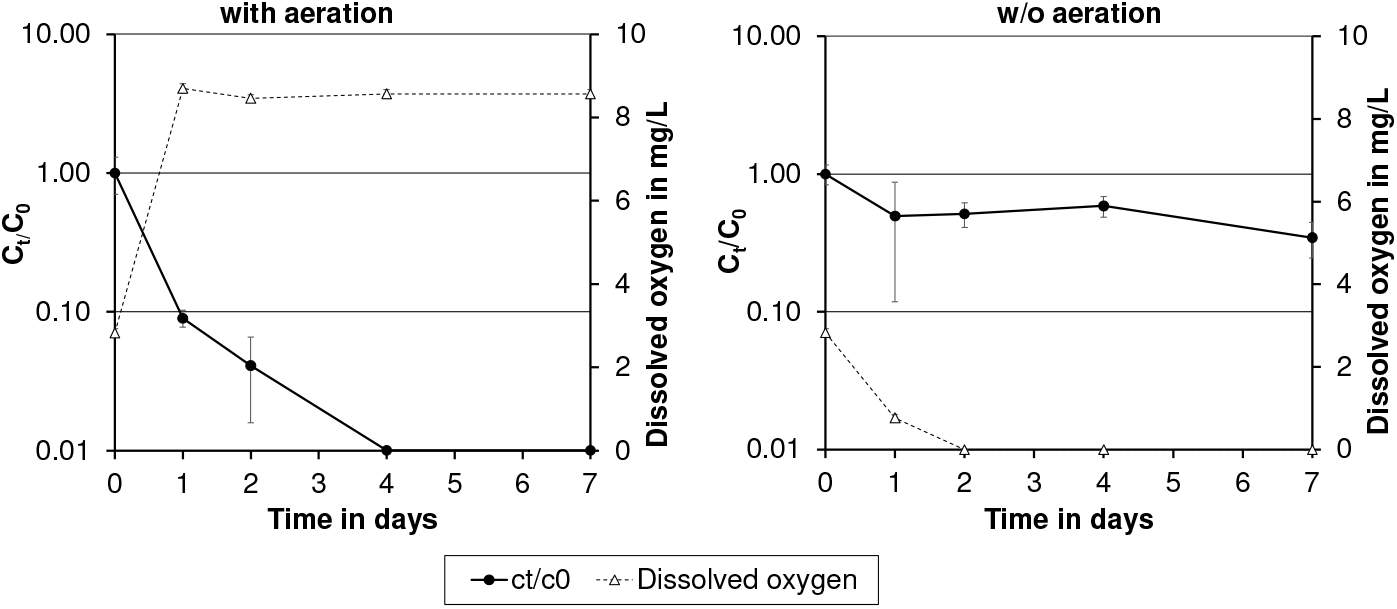
Decay of SARS-CoV-2 biomarkers (as means of ORF, E, RdRP2 and NSP3 copy numbers) and oxygen concentration over time in wastewater samples purged with air (left) or nitrogen (right). Errror bars represent standard deviations for the three independent test bottles (n = 3).

The virus stability studies published so far have mostly used other human or animal pathogenic coronaviruses, showing an influence of temperature, pH and matrix on the decay of coronaviruses in several water types (Foladori et al., 2020). At room temperature, coronaviruses suspended in tap water, primary WWTP effluent or untreated and autoclaved wastewater showed 99.9% reduction within 2–5 days (Ahmed et al., 2020; Gundy et al., 2009). In contrast, animal coronaviruses such as transmissible gastroenteritis virus (TGEV) and mouse hepatitis virus (MHV) showed a higher persistence in pasteurised settled sewage at room temperature (Casanova et al., 2009); times for a 99% reduction were 7 days for MHV and 9 days for TGEV (Casanova et al., 2009). However, oxygen concentration has not been measured in these studies.

It has been known for a long time that the oxygen concentration can be an important factor influencing virus survival. Scheuerman et al. (1991) have studied the effects of temperature and DO on the persistence of enteric viruses in sludge (polio 1, coxsackie B3 and echo 1, and rotavirus SA-11) under laboratory conditions. The inactivation rates under aerobic conditions were significantly higher than those under anaerobic conditions (−0.77 log_10_/day vss -0.33 log_10_/day) (Scheuerman et al., 1991). Also in groundwater studies, the presence of oxygen accelerated enteric viruses decay (Gordon and Toze, 2003; Sidhu and Toze, 2012).

## 4. Conclusions

Within this study, a robust method for the detection of SARS-CoV-2 RNA in wastewater was established. This detection methodology is based on PEG precipitation followed by automated nucleic acid extraction and the detection of different target genes by RT-ddPCR. The following findings were obtained:

- Comparison of ddPCR results for seven different primer and probe assays showed that E, ORF, RdRP2 and NSP9 were suitable for wastewater monitoring and yielded similar results. The N1, RdRP1 and NSP3 assays clearly deviated from the average of the other primer and probe combinations. Based on these results and the high mutation potential of coronaviruses, we recommend the parallel detection of at least two, preferably three or more, target sequences to increase the robustness of the method.
- Comparison of the wastewater monitoring results for the city of Karlsruhe with reported COVID-19-infection numbers confirmed the potential of WBE for the early detection of trends in the incidence of infection within a catchment area.
- With PCR-based wastewater monitoring also the spread of VoCs in a catchment area can be monitored, making it a fast and cost-effective alternative to NGS-based approaches.
- SARS-CoV-2 RNA in wastewater showed low decay under anaerobic conditions, and fast decay under aerobic conditions. These findings are relevant for a better understanding of biomarker stability in sewer systems, and storage of wastewater samples.

The data obtained from the wastewater monitoring were made available to the Covid-19 Task Force of the city of Karlsruhe and used as an additional tool for pandemic management.

## Supporting information

Raw data

## Data Availability

Raw data is uploaded next to SI.

## Acknowledgements

This study was financially supported by the German Federal Ministry of Education and Research as part of the funding program Sustainable Water Management (NaWaM-RiSKWa) (Biomarker, grant number 02WRS1557). We thank the City Civil Engineering Office of the city of Karlsruhe (Tiefbauamt der Stadt Karlsruhe), especially Martin Kissel, Albrecht Dörr, Dr. Gaby Morlock und Stephen Kemper. We also thank Dr. Marion Woermann und Carmen Kraffert for their assistance with wastewater monitoring and experimental work.

---

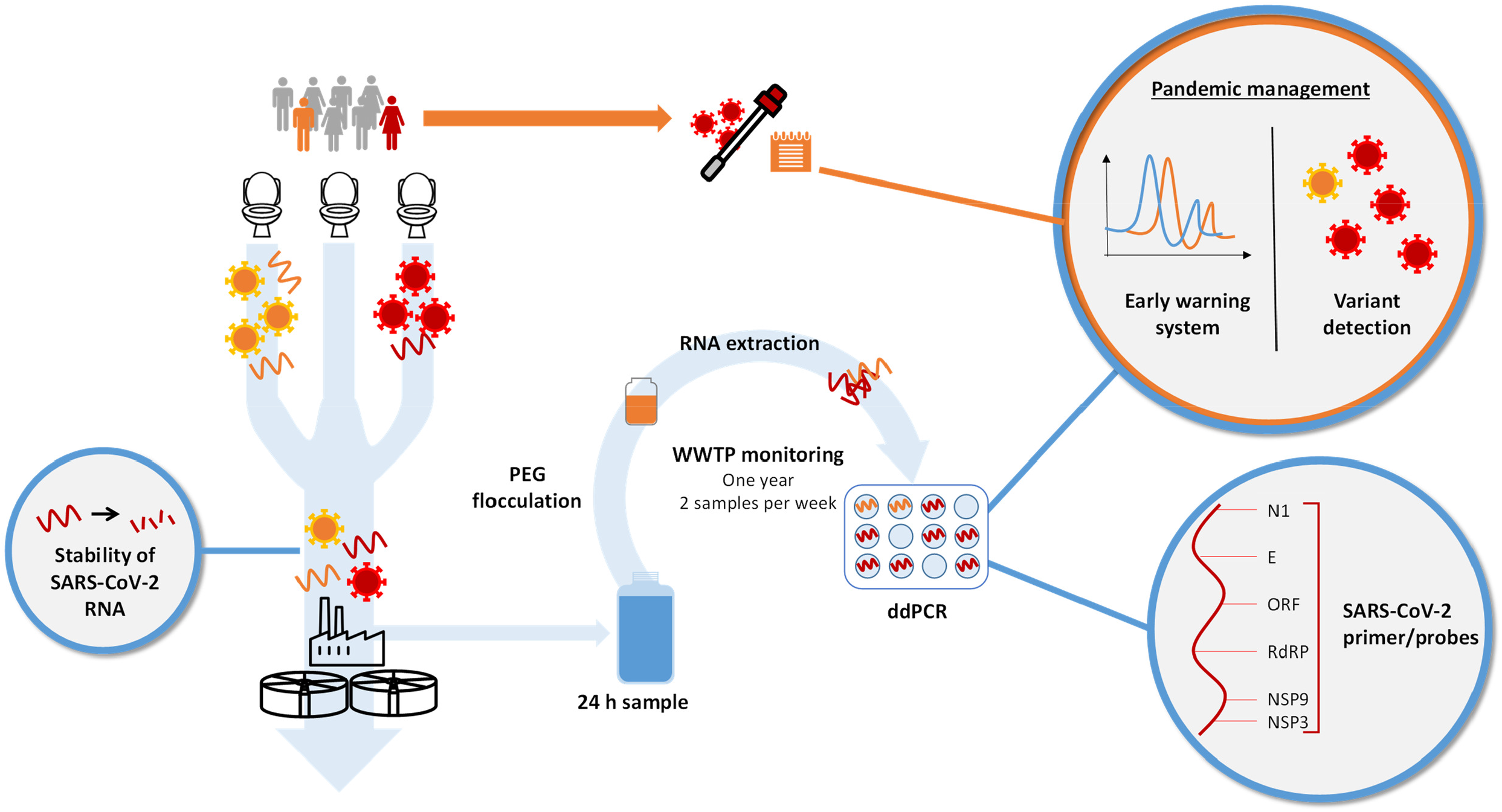

