## Supplementary material for "SARS-CoV-2 wastewater surveillance in Germany: long-term PCR monitoring, suitability of primer/probe combinations and biomarker stability": SI

Infection value shift in days

| | 0 | 1 | 2 | 3 | 4 | 5 | 6 | 7 | 8 | 9 | 10 | 11 | 12 | 13 | 14 | 15 | 16 | 17 | 18 | 19 | 20 | 21 |
|---|---|---|---|---|---|---|---|---|---|---|---|---|---|---|---|---|---|---|---|---|---|---|
| $r^2$ | 0.44 | 0.49 | 0.56 | 0.60 | 0.63 | 0.67 | 0.71 | 0.75 | 0.77 | 0.80 | 0.81 | 0.83 | 0.85 | 0.86 | 0.87 | 0.88 | 0.89 | 0.88 | 0.87 | 0.85 | 0.84 | 0.83 |

**Fig. S1.** Correlation coefficients ($r^2$) for infection numbers and biomarker concentrations with different time shifts from 0 to 21 days for the infection numbers. P values with Bonferroni-correction were below 0.001 for all correlations.

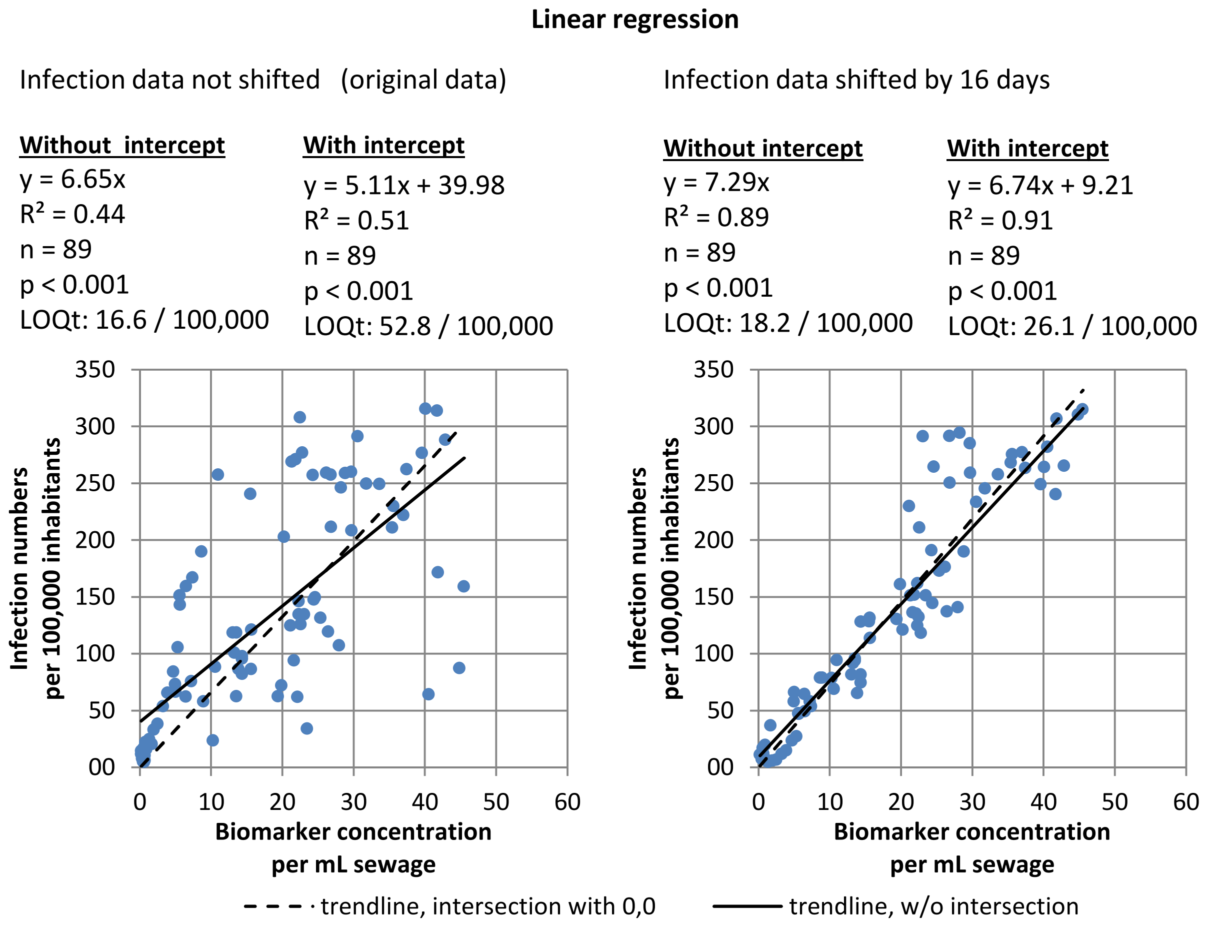


**Fig. S2.** Linear regression for infection data and ddPCR values (E and ORF gene as sliding average of three values). Left: original data, right: infection numbers were time-shifted by 16 days. Statistical analysis above each plot for each trend line with and without intercept. P values with Bonferroni-correction were below 0.001 for all correlations.

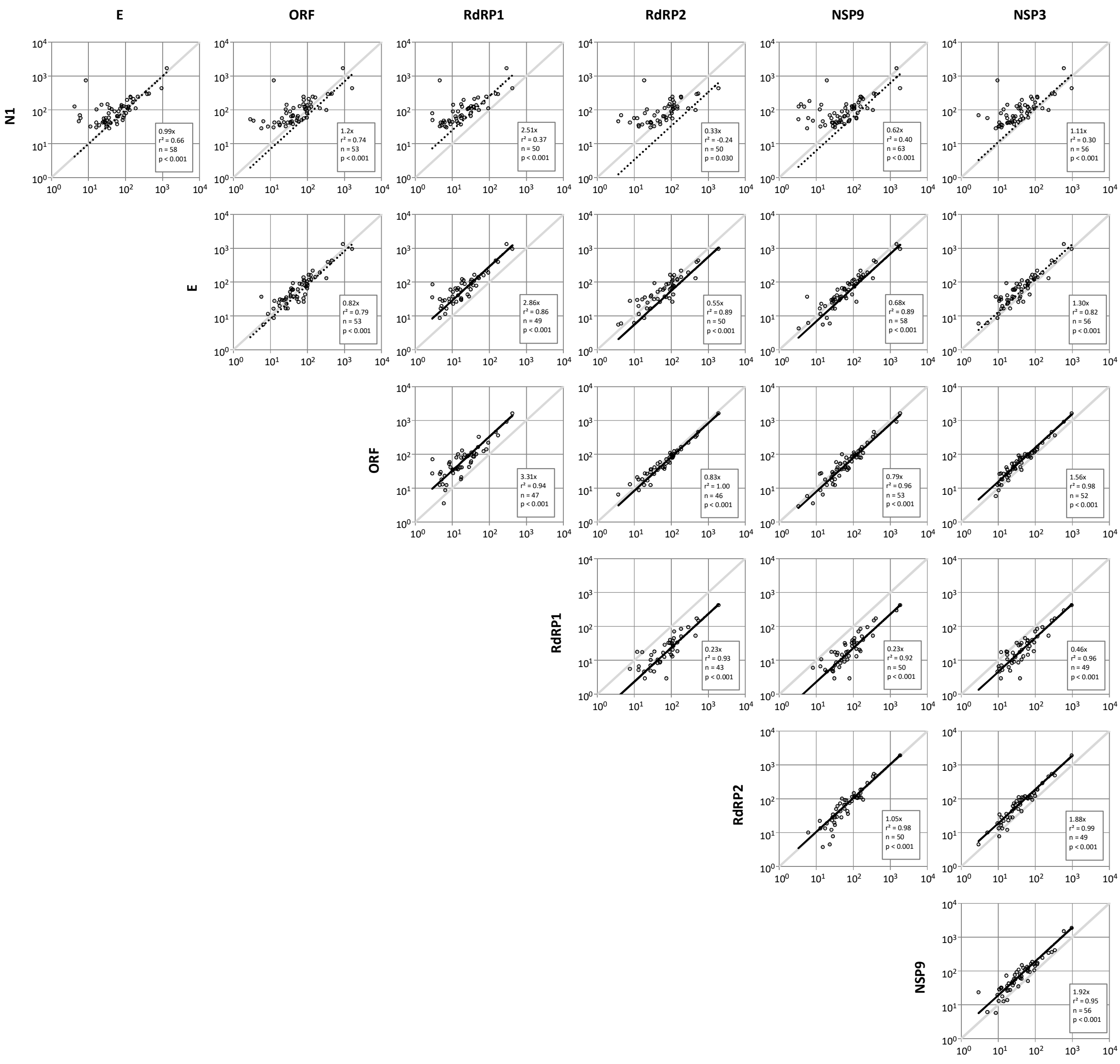


**Fig. S3.** Linear regressions of seven different primer/probe assays using a selection of 64 samples. Charts show gene copies per mL wastewater.

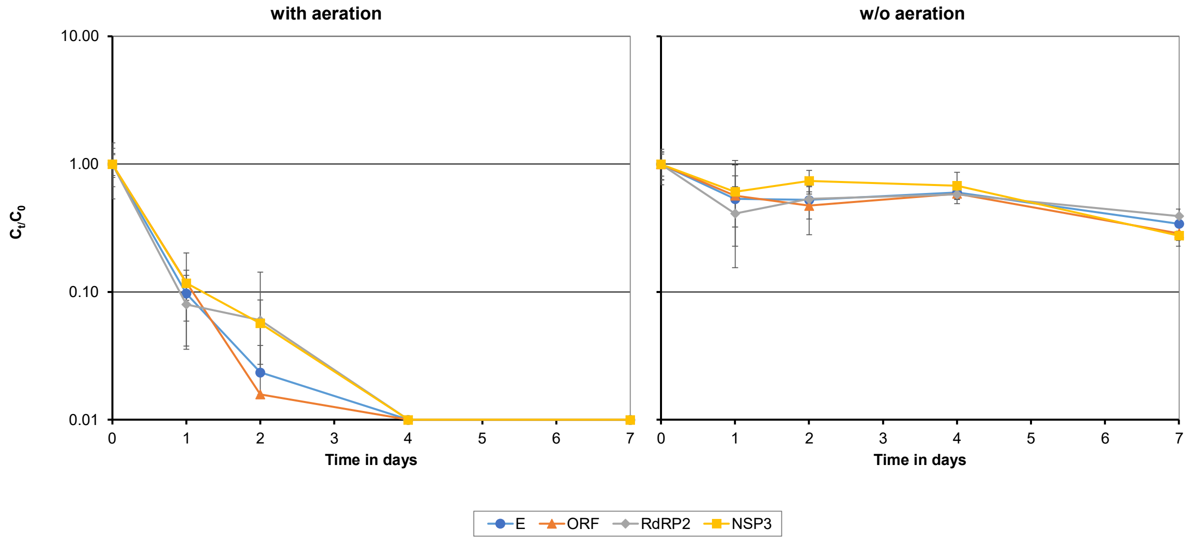


**Fig. S4.** Decay of SARS-CoV-2 specific targets over time in wastewater samples aerated with air (left) or nitrogen (right). Errror bars represent standard deviations for the three independent test bottles (n = 3).
